# Limited evidence for validity and reliability of non-navigated low and high frequency rTMS over the motor cortex

**DOI:** 10.1101/2023.01.24.23284951

**Authors:** Kilian Prei, Carolina Kanig, Mirja Osnabrügge, Berthold Langguth, Wolfgang Mack, Mohamed Abdelnaim, Martin Schecklmann, Stefan Schoisswohl

## Abstract

The neuromodulatory effects of repetitive transcranial magnetic stimulation (rTMS) are often described as inhibiting for low frequency and facilitating for high frequency protocols, leading to the lofi-hife heuristic. However, the data basis for this is inconsistent and reliability of rTMS is barely evaluated. The present study examines the validity of this lofi-hife heuristic at group and single subject level and the reliability of rTMS in a non-navigated setting close to clinical application. In 30 healthy participants, 1 Hz and 20 Hz rTMS were each administered twice over the left motor cortex resulting in four sessions/participant. Motor evoked potentials (MEPs) were measured before and after each session. Reliability measures were intraclass and Pearson’s correlation coefficient (ICC and *r*). The heuristic was not evident at group level. At single-subject level four participants responded with heuristic-conform changes, i.e., concomitant decreases for 1 Hz and increases for 20 Hz sessions. ICCs and *r* were low to moderate. Within subgroups of less confounded measures, we found good *r* values for 20 Hz rTMS. Results demonstrate high inter- and intraindividual variability of rTMS questioning the lofi-hife heuristic. Methodological improvements for the usage of rTMS might help to increase validity and reliability of non-invasive brain stimulation.

## Introduction

Transcranial magnetic stimulation (TMS) is a non-invasive brain stimulation technique whereby electromagnetic pulses are administered to the skull. The change of magnetic flux can lead to the depolarization of underlying cortical neurons [1,2]. Single TMS pulses applied over the primary motor cortex (M1) can elicit contralateral peripheral motor responses termed motor evoked potentials (MEPs), which are measured by electromyography (EMG) [3]. MEPs are typically used as a parameter of cortical excitability [4]. The application of TMS pulses in a rhythmic manner with specific frequencies is referred to as repetitive transcranial magnetic stimulation (rTMS) [5] and is capable to induce changes in cortical excitability, that can persist beyond stimulation offset [6,7]. It is assumed that the changes in cortical excitability (i.e. neuroplasticity) are frequency dependent: low frequency rTMS (≤ 1 Hz) induces inhibitory effects whereas high frequency rTMS (≥ 5 Hz) evokes excitatory effects [8– 12], hereinafter referred to as lofi-hife heuristic (**lo**w **f**requency **i**nhibitory - **hi**gh **f**requency **e**xcitatory).

Several study results were incompatible to the lofi-hife heuristic questioning its validity. Various researchers did neither realize inhibiting effects with low frequency protocols (e.g., 1 Hz) [13–16] nor facilitatory effects with high frequency protocols (e.g., 20 Hz) [11,16,17].

In line with limited evidence for the validity of lofi-hife heuristic, examinations of reliability of rTMS are scarce. Until now, the reproducibility of commonly used 1 and 20 Hz rTMS protocols has been investigated in only 5 studies [9,11,14,18,19]. Four of them only compared MEP amplitudes or MEP area under the curve between sessions using rather small sample sizes (*n* = 4 - 10) and did not report any measure of reliability [9,14,18,19]. The sole experiment which calculated Pearson’s correlation coefficients (*r*) was conducted by Maeda et al. [11] (*n* = 20). Maeda and colleagues demonstrated correlations between low (*r* = .266 for 1 Hz and *r* = .260 for 10 Hz; *p* > .050) and moderate (*r* = .543 for 20 Hz; *p* = .013) effect size. The reliability of other TMS-based neuromodulatory methods like intermittent and continuous theta-burst stimulation (TBS) is classified as low to moderate as well [20,21].

Given that there is high within- and between-subject variability [22–24] and that there is a limited number of studies on the reliability of rTMS, the aim of the present work is to investigate the day-to-day retest reliability of a low and a high frequency rTMS protocol (1 Hz, 20 Hz) on group and single-subject level as it is commonly used in experimental and clinical environments. We therefore aimed to keep our experimental setup as close to clinical application as possible. Additionally, we tested whether we could reproduce lofi-hife heuristic-conform effects.

## Methods

### Sample characteristics

The analyzed sample consisted of 30 healthy volunteers (22 female; age range: 19 – 29 years, *M* = 22.90, *SD* = 2.75). The included participants had an estimated average intelligence quotient of 110.2 (*SD* = 8.8) according to the Multiple Selection Vocabulary Test [25], scores higher than 33.3 in the Edinburgh Handedness Inventory (*M* = 85.0, *SD* = 16.8; only left-handed: -100, only right-handed: +100) [26] and less than 20 points (cut-off for clinic relevant depression) in the Major Depression Inventory (MDI; *M* = 5.3, *SD* = 3.3) [27]. Starting with a sample of 41 participants, two were excluded due to indications of depression via MDI and Structured Clinical Interview for DSM-IV Axis I Disorders [28], assessed by a trained clinical professional. Moreover, nine people with a resting motor threshold (RMT) above 55% maximum stimulator output were excluded in order to prevent overheating of the passively cooled TMS coil and increase the stimulation tolerability for the volunteers. The included participants had no regular intake of medications apart from contraceptives.

The study protocol, which is according to the Declaration of Helsinki, was reviewed and approved by the local ethics committee of the University of Regensburg (ethical approval number: 16-101-0305). All participants gave their informed consent before the start of the experiment.

### Study procedure

The entire experiment consisted of a pre-experimental session in which the motor hotspot and RMT of the participants were determined as well as the questionnaires completed, which was followed by the main experiment with four consecutive rTMS sessions (**Figure 1**). All experiments were conducted by the same investigator at the Centre for Neuromodulation at the University of Regensburg, Germany.

**Figure 1.** Study procedure. The pre-experiment included only a resting motor threshold (RMT) determination. Each day of the main experiment comprised a RMT determination, a pre-measurement of cortical excitability with 132 motor evoked potentials (MEPs), a protocol of repetitive transcranial magnetic stimulation (rTMS) (1800 pulses, 1 Hz or 20 Hz) and a post-measurement identical to the pre-measurement. To analyze the time-course after rTMS, the 132 MEPs post-measurement were divided up into quarters of 33 MEPs (Q1-Q4).

In the pre-experiment, we searched the hotspot where TMS pulses elicited the highest and most stable MEPs. To do this, we started from the C3 electrode position (10-20 electroencephalography system) with an orientation at about 45° of the TMS coil in posterior-anterior direction to the sagittal midline of the head and the handle pointing backwards. The hotspot and landmarks for placing were marked on a blank cap to ensure a coil positioning as constant as possible in subsequent stimulation sessions. RMT was defined as five out of ten TMS pulses inducing a MEP with a peak-to-peak amplitude of at least 50 µV [29,30].

The main experiment took place over four days from Monday through Friday. In four participants one session was postponed to a later appointment due to scheduling conflicts. On each day one study session was conducted. The four sessions consisted of two identical stimulation sessions for 1 Hz and 20 Hz rTMS with both frequencies in alternating order (cf. **Figure 1**). The protocol used first was set via balanced randomization. Each session was conducted at the same time of day (either always 8 am, 10 am, 12 noon or 2 pm) to prevent intrasubject effects due to the circadian rhythm [31]. The participants were required to abstain from caffeine, alcohol or medication excluding contraceptives, and to maintain their current sleep rhythm throughout the experiment.

At the beginning of each session, the caps were put on based on the landmarks followed by motor hotspot and RMT determination, which started from the position marked in the pre-experiment. Potential changes in hotspot position were marked on the cap. The participants were asked to lie as still and relaxed as possible with no explicit attention focus, but to keep their head and right hand in a steady position, which was monitored visually. In addition, we used a vacuum pillow and a coil holder to ensure that the head of the participant remained in its position relative to the coil as good as possible. Coil position and EMG data were observed during the experiment and in case of coil-slipping from the marked hotspot or the absence of EMG responses on TMS pulses for more than a minute the coil position was corrected and documented as “coil correction” within a session.

### Repetitive transcranial magnetic stimulation (rTMS)

For TMS and the subsequent rTMS sessions we used a MAGPro X100 stimulator as well as a figure-of-eight coils of the model MCF-B65 (all Medtronics Plc, Ireland). For each participant, the same coil was used throughout the entire experiment.

All rTMS-protocols included 1800 stimuli administered at 110% of RMT on the individual motor hotspot. During the low frequency sessions, the stimulation was performed at 1 Hz for 30 min continuously. During the high frequency sessions, 45 trains of 40 stimuli were given at 20 Hz with inter-train intervals of 28 s, resulting in a total rTMS time of 22 min. The procedures matched those of a former experiment [32].

### Motor evoked potentials (MEPs)

Before and directly after rTMS, we performed MEP measurements consisting of 132 TMS pulses applied at 110% RMT in the same manner on all four days. The stimuli were presented with a jittered interstimulus interval of 10 ± 2 s via Presentation (Version 20.1, Neurobehavioral Systems Inc., United States). This resulted in a mean duration of 22 min per MEP measurement.

MEPs were recorded via surface electrodes (Ag/AgCl) from the right first dorsal interosseus muscle (FDI) in belly-tendon montage. The ground and reference electrodes were fixed on both styloid processes. The signal was amplified by a 16-channel amplifier (V-Amp, Brain Products GmbH, Germany), processed with Brain Vision Recorder V-Amp Edition (Version 1.10, Brain Products GmbH, Germany) and analyzed offline with Brain Vision Analyzer (Version 1.05.0005, Brain Products GmbH, Germany). The sampling rate was 500 Hz for the first four and 2000 Hz for the remaining 26 participants. A zero phase Butterworth filter was applied (1 Hz cutoff, notch filter at 50 Hz). The stimulus interval was defined from 10 ms before to 60 ms after the stimulus. MEP amplitudes were calculated peak-to-peak. For further analysis we considered segments with a prominent MEP pattern (biphasic signal with first a minimum, followed by a maximum; visually distinct from the baseline) as valid. Invalid segments were excluded from further analyses. While other researchers excluded all MEPs with amplitudes below 50 µV [33] or outside 2.5 SDs from average amplitude [20,21], we wanted to include these MEPs to avoid losing MEP changes from before to after rTMS.

### Statistical analysis

#### Preprocessing

We used SPSS Version 25.0 (IBM Corp., USA) for statistical analyses. The significance level was 5% for all computations. We calculated the mean (*M*) and, to include a more resistant measure against outliers, the median (*Md*) of the entire MEP measurement, i.e., 132 segments, and the quarters per measurement (Q1-Q4; 33 consecutive measures each).

To ensure comparability of MEP responses across measurements, we were interested in the amount of coil corrections and invalid trials per measure. Overall, 17.5% of the segments needed to be excluded according to the above-mentioned criteria and in 19.17% of the pre and post measurements, the coil needed to be adjusted. With three subsamples of participants (*n* = 11 participants without coil correction; *n* = 9 participants with at least 75% valid MEPs in every condition; *n* = 7 participants with no coil correction and > 75% valid MEPs) we repeated the analyses on a group level to see if this preselection leads to more prominent effects or improved reliability.

#### Effects of rTMS: lofi-hife heuristic

2 (“frequency”; 1 Hz vs 20 Hz) x 2 (“session”; first vs second) x 2 (“within-session time”; pre vs. post rTMS) x 4 (“quarter”; Q1-Q4) repeated measurement ANOVAs were performed for *M* and *Md*. These analyses aimed to assess the validity of the lofi-hife heuristic by looking for frequency-specific rTMS-induced changes (“frequency” x “within-session time”) and whether these interactions were stable during the follow-up periods and between the sessions (influence of the factors “quarter” and “session”). Post-hoc paired t-tests were conducted in case of main or interaction effects with *p*-values adjusted by false discovery rate (FDR) due to multiple testing [34,35].

At single-subject level, we calculated the number of participants whose MEP amplitudes were either significantly (t-test) or descriptively higher, lower or unchanged after the respective rTMS intervention in comparison to before. To further elaborate the individual day-to-day congruence of MEP response to the different frequencies, we conducted Chi-squared tests with the contingency tables of both sessions for 1 Hz and 20 Hz stimulation. Thus, we were able to derive information about the stability of rTMS effects and hints to reliability at single-subject level.

#### Reliability

Reliability group measures were the intraclass correlation coefficient (ICC) and Pearson’s correlation coefficient (*r*). Since we were interested in rTMS induced effects, we used the differences from post (for the whole measurement and the four quarters) to pre (for the whole measurement) of *M* and *Md* to adjust for the baseline activity in each session. We calculated two-way mixed effect ICCs with general agreement and *r* for each dependent variable (*M* and *Md*) for the whole measurement as well as the separate quarters between the 2 sessions of both 1 Hz and 20 Hz rTMS protocols. ICC and *r* results were compared to zero, the p-values were FDR adjusted within the group of five values (whole measurement and four quarters).

## Results

### Effects of rTMS: lofi-hife heuristic

According to the lofi-hife heuristic, we expected an interaction effect of “within-session time” and “frequency” in the 2×2×2×4 - ANOVAs for *M* and *Md* (increase of MEPs from pre to post for 20 Hz and decrease for 1 Hz), but this interaction remained non-significant (*M*: *F*(1, 29) < 1, *p* = .533, η ^2^_*p*_ = .014; *Md*: *F*(1, 29) < 1, *p* = .453, η ^2^_*p*_ = .020) (cf. **Figure 2)**. Interactions of “within-session time” and “frequency” with “quarters” or “session” were not found as well (*M*: *F*(2.16, 62.66) < 1, *p* = .407, η ^2^_*p*_ = .031 and *F*(1, 29) = 1.458, *p* = .237, η^2^_*p*_ = .048; *Md*: *F*(1.66, 48.13) < 1, *p* = .614, η ^2^_*p*_ = .022 and *F*(1, 29) = 1.846, *p* = .185, partial η ^2^_*p*_ = .060)). The Greenhouse– Geisser adjustment was used for the “within-session time” x “frequency” x “quarters” interaction of *M* and *Md* to correct for violations of sphericity. Within ANOVAs of the three subsamples (at least 75% valid MEPs, no coil correction and both), none of the expected interactions was evident.

**Figure 2.** “Frequency” x “ within-session time” interaction vs. expectation. While **a** depicts our actual findings, **b** shows the expectation according to the heuristic (interaction with increase of MEP amplitudes after 20 Hz and decrease of MEP amplitudes after 1 Hz rTMS) schematically. Error bars represent standard error. *M* = Mean.

We also analyzed rTMS-induced changes on a single-subject level. We inspected the changes from pre to post for each participant on a descriptive level and also with a t-test. Based on the heuristic, a decrease for 1 Hz and an increase for 20 Hz was expected. 2 out of 30 participants showed a significant decrease of MEPs from pre to post 1 Hz rTMS in both sessions. One participant had significant MEP increases after both 20 Hz sessions, this participant was the only person in the sample who responded in all 4 sessions with significant changes according to the lofi-hife heuristic. Chi-square tests of independence showed no association between session 1 and 2 for 1 Hz (χ ^2^ (4) = 8.914, *p* = .062) and 20 Hz (χ ^2^ (4) = 2.066, *p* = .755) (see **Table 1**). According to descriptive inspection of data, 14 out of 30 participants showed a decrease of MEPs from pre to post 1 Hz rTMS in both sessions. Likewise, after 20 Hz rTMS stimulation, 9 out of 30 participants had higher MEPs in both sessions. Four participants showed decreases in MEPs due to 1 Hz and increases due to 20 Hz concomitantly. Based on Chi-squared tests of independence, changes after rTMS on single-subject level were significantly associated for 1 Hz rTMS between both sessions with eight participants showing increases and 14 decreases (see **Table 2**) (χ ^2^ (1) = 6.696, *p* = .019). For 20 Hz (χ ^2^ (1) < 1, *p* = .713) there was no association between session 1 and 2.

**Table 1.**
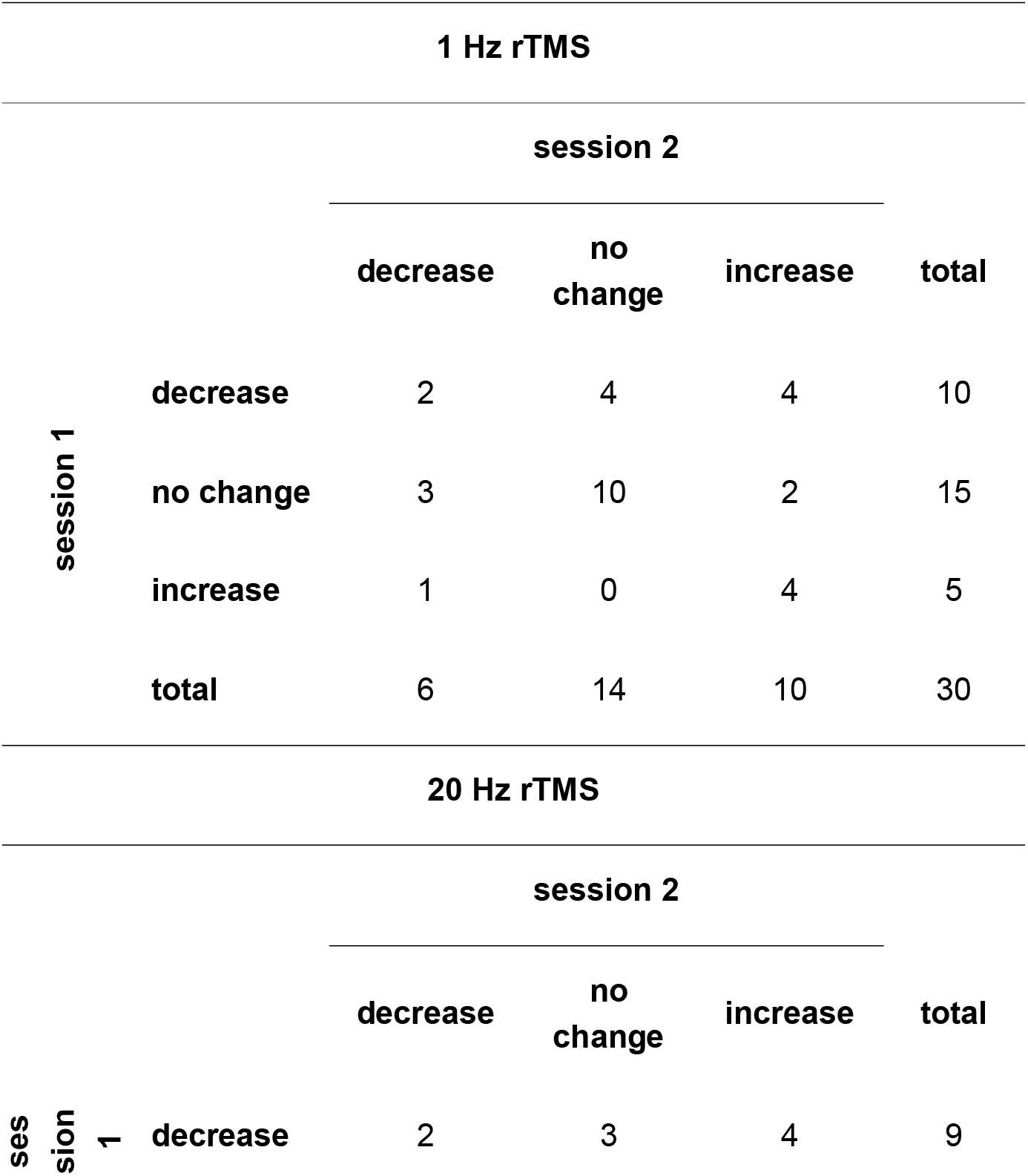
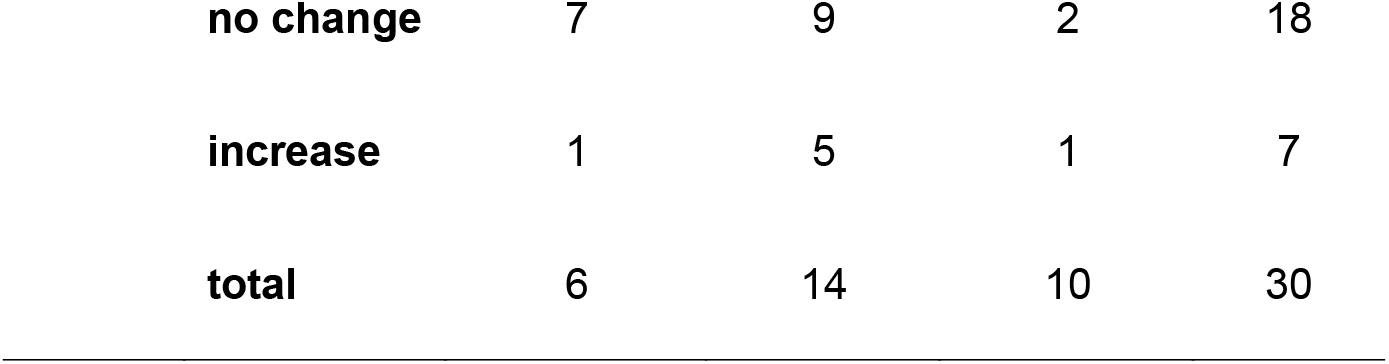
Contingency tables for significant changes of the amplitudes of motor evoked potentials (MEPs) from pre to post for the individual participants *Notes*. Significant changes refer to the difference in the MEP amplitude of from pre to post which is significant in a paired t-test.

| 1 Hz rTMS |  |  |  |  |  |
| --- | --- | --- | --- | --- | --- |
|  |  | session 2 |  |  |  |
|  |  | decrease | no change | increase | total |
| session 1 | decrease | 2 | 4 | 4 | 10 |
|  | no change | 3 | 10 | 2 | 15 |
|  | increase | 1 | 0 | 4 | 5 |
|  | total | 6 | 14 | 10 | 30 |
| 20 Hz rTMS |  |  |  |  |  |
|  |  | session 2 |  |  |  |
|  |  | decrease | no change | increase | total |
| session 1 | decrease | 2 | 3 | 4 | 9 |

|  |  |  |  |  |
| --- | --- | --- | --- | --- |
| **no change** | 7 | 9 | 2 | 18 |
| **increase** | 1 | 5 | 1 | 7 |
| **total** | 6 | 14 | 10 | 30 |

**Table 2.**
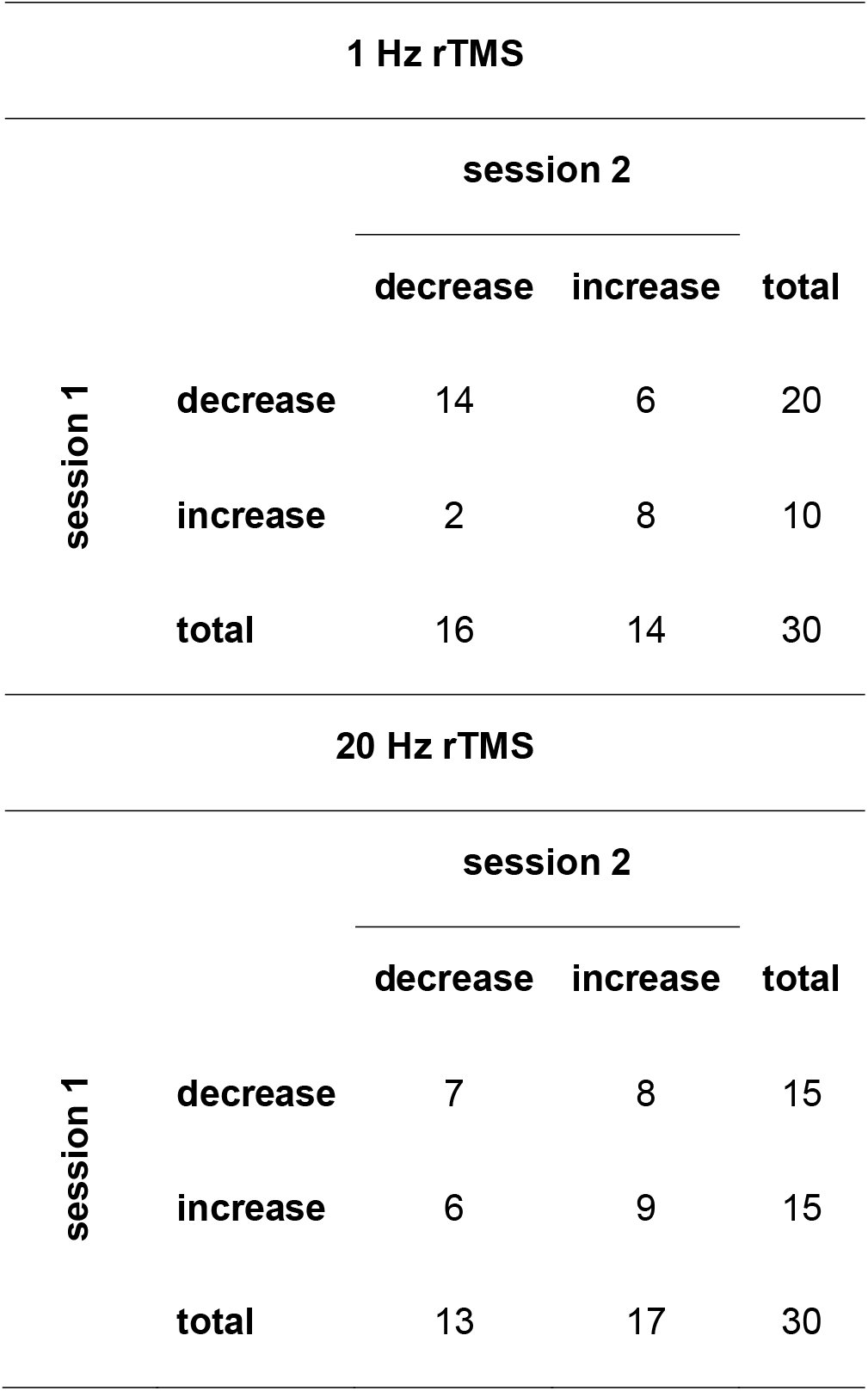
Contingency tables for descriptive changes of the amplitudes of motor evoked potentials (MEPs) from pre to post for the individual participants *Notes*. Descriptive changes refer to a greater-than/smaller-than relation between the means of MEP amplitudes between pre and post.

| **1 Hz rTMS** |  |  |  |  |
| --- | --- | --- | --- | --- |
|  |  | **session 2** |  |  |
|  |  | **decrease** | **increase** | **total** |
| **session 1** | **decrease** | 14 | 6 | 20 |
|  | **increase** | 2 | 8 | 10 |
|  | **total** | 16 | 14 | 30 |
| **20 Hz rTMS** |  |  |  |  |
|  |  | **session 2** |  |  |
|  |  | **decrease** | **increase** | **total** |
| **session 1** | **decrease** | 7 | 8 | 15 |
|  | **increase** | 6 | 9 | 15 |
|  | **total** | 13 | 17 | 30 |

### Test-retest reliability

ICC and *r* for *M* and *Md* classified the test-retest reliability of rTMS effects for low and high frequency protocols mostly as poor (ICC < .5; *r* < .3). After FDR adjustment, no correlations (ICC and *r)* were identified, that are significantly divergent of zero, neither for the whole measurement nor for the quarters. There were no significant reliability differences over time (i.e., over the course of the 132 pulses after the rTMS), but descriptively the lowest reliability values occurred for both 1 and 20 Hz. ICC and *r* for the whole sample are depicted in **Figure 3**.

**Figure 3.** Intraclass correlation coefficient (ICC) (**a**) and Pearson’s correlation coefficient (*r*) (**b**) between session 1 and 2. Calculations were made for the whole measurement as well as for the quarters: quarter 1 (Q1), quarter 2 (Q2), quarter 3 (Q3), quarter 4 (Q4). Error bars represent the upper/lower bound of the 95% confidence interval. Colors represent the reliability/effect size areas as trivial (darker red), poor/low (lighter red), medium/moderate (yellow), good/large (lighter green) or excellent (darker green). Numerical values of ICC and *r* are listed in Supplementary Table S1. and *M* = mean; *Md* = median.

For the three subsamples (at least 75% valid MEPs, no coil correction and both), in the 20 Hz sessions, ICCs ranged up to .67 and *r* up to .77 and in the 1 Hz sessions up to .22 and .12, respectively. We found significant ICC measures after FDR correction in the ICCs of the subsample with at least 75% valid MEPs for *M* in the fourth quarter of 20 Hz stimulation (ICC = .667, *p*_FDR_ = .049) and for *Md* in the second quarter of 20 Hz stimulation (ICC = .619, *p*_FDR_ = .042). No other significant correlations were detected for the three subsamples.

## Discussion

The present paper studies the validity of the lofi-hife heuristic and the day-to-day reliability of rTMS effects in a non-neuronavigated setup on group and single-subject level. In sum, we found no evidence for the lofi-hife heuristic. Reliability measures stayed within a low to medium range.

### Validity of the lofi-hife heuristic

We could not reproduce the statements of the lofi-hife heuristic. At group level, the expected interaction of “frequency” and “within-session time” in *M* and *Md* where 1 Hz rTMS leads to inhibition and 20 Hz to facilitation of cortical excitability did not occur. An influence of the factor “quarter” on this interaction which might hint to a fading of the effects [36] also failed significance. Stronger rTMS effects on repeated measures have been reported before [9,37], but in our case no corresponding interaction with the “session” factor was shown.

The analyses of subsamples with no coil corrections, at least 75% valid MEPs or both yielded no heuristic-conform effects as well. Still, the statistical power of these analyses with fewer participants is lower compared to the whole sample, which is why present effects might be undetected yet.

At single-subject level, 27% of all 1 Hz measurements and 28% of all 20 Hz measurements showed significant heuristic-conform neuroplastic changes (for descriptive differences 60% and 53%, respectively). Therefore, it was found that almost three quarters of the measurements were not in line with the heuristic and showed either no changes or even changes opposite to the heuristic. Only one participant (for descriptive differences four participants) reacted according to the heuristic in all four measurements.

Overall, our study found no evidence for the lofi-hife heuristic at group and single subject level. The literature reveals ambiguous results about the effect of rTMS over the motor cortex on cortical excitability [10], which is unsurprising considering the impact of variability [23,24,38]. Many factors such as the participants’ intrinsic state [39], age [40] or medication [41] can influence the effects of rTMS and not all factors may have been identified yet [22]. Therefore, variability of outcome measures is a common challenge in the interpretation of dependent variables.

Closely related to the question of the validity of the heuristic and the challenge due to variability is the question of the reliability of the rTMS effects. Solely looking at the direction of the descriptive MEP changes from pre to post, a significant relation between the 2 sessions of 1 Hz rTMS emerged. Notably, this effect was caused not only by the 47% of participants who responded in conformity with the lofi-hife heuristic, but also by the 27% of participants who responded excitatory. Stable effects which are independent of the heuristic can therefore still occur.

### Test-retest reliability

In our setting, test-retest reliability at group level was low to moderate for *Md* and *Md* for both ICC [42] and *r* [43]. Reliability measures for *Md* were descriptively slightly larger than for *M*. Therefore, the inclusion of outliers in the analyses may lower the correlation coefficients. While a standardized method of dealing with outliers is missing [44], methods such as using the *Md* or excluding amplitudes outside of 2.5 SD ± *M* could lead to more stable results. However, it remains unclear whether these methods are truly suited for representing cortical excitation better than evaluations that include the outliers.

The magnitude of reliability measured by *r* for 1 Hz rTMS is similar to Maeda et al. [11], where descriptively higher values for 20 Hz (*r* = .543) were demonstrated. Stimulation parameters differed from our setup, thus hampering comparability. Maeda et al. used a subthreshold stimulation intensity, a shorter protocol (4 min and 240 stimuli vs. 22 – 30 min and 1800 stimuli) and pre- and post-measurements were based on fewer MEPs (10 vs. 132 trials). Previous research suggests that our setting with a higher stimulation intensity and protocols with more pulses would result in more reliable effects [45]. Nevertheless, we did not achieve better reliability than Maeda et al. Ten MEPs cover only the first 3 min after rTMS, comparable to Q1 (0 - 5.5 min) of our study, where we achieved descriptively the lowest reliability measures. Looking at TBS protocols, the lowest reliability was observed directly after the intervention at 5 and 10 min as well. Better reliability was achieved in the second half of the recording (between 30 and 60 min post continuous TBS) [21].

In the subgroups, we expected better reliability measures due to the exclusion of the possible confounding factors coil corrections and (high number of) invalid MEPs. Whereas no such tendency was seen for 1 Hz, 20 Hz showed improved values for ICC and *r*. For the group with at least 75% valid MEPs, two ICCs differed significantly from zero. Thus, the role of controlling the coil position and achieving valid MEPs may be more important for 20 Hz protocols than for 1 Hz protocols. Discomfort, pain and muscle twitching caused by high frequency rTMS is more often reported than in case of low frequency stimulation [46] and make arousal and movements more likely. This can lead to coil slipping and invalid MEPs and thus needs to be controlled.

While our study revealed at best moderate reliability for rTMS induced MEP changes, the true “intrinsic” reliability of rTMS effects on cortical excitability remains unclear. Many confounding factors in rTMS-MEP setups can lead to variability and therefore may lower the reliability. An overview of these factors is provided by various reviews [23,24,44,47]. Our study design tried to incorporate many recommendations such as the exclusion of drug intake, the constant time of day for each participant, the use of more than 30 trials to estimate the MEP amplitude and the sample size [44]. Other factors such as genetic pre-screenings [48] or the use of closed-loop systems to evoke TMS pulses in similar brain-states [49] were not incorporated in our setting, because we wanted to estimate the reliability of a setting that corresponds to everyday use of rTMS in a clinical or research context. An interesting question for future research concerns the maximal realizable reliability of rTMS and which methodological arrangements are optimal to achieve this. The descriptively higher reliability in the subgroups for 20 Hz indicates potential for improvement.

Thus, more valid MEPs may improve reliability. Input–output slope curve estimation is an approach to determine the optimal stimulation intensity with low risk for ceiling or floor effects [50]. At the moment, RMT determination according to Rossini and Rothwell [29,30] is better investigated, commonly used and showed at least good reliability [51,52]. But with I-O curves stimulation intensity can be tuned with respect to individual limits.

Furthermore, neuronavigation and can ensure a stable position of the coil in relation to the cortical target [53,54]. While some studies postulate increased effects and reliability of rTMS through the use of neuronavigation [55–57], others show no further MEP reliability improvement [58,59] or no benefit in treatment [60,61]. In addition to neuronavigation, collaborative robots (cobots) may presumably further reduce the positioning error [62]. However, cobots and neuronavigation systems are still costly and not present in most labs and hospitals. Open-source projects for cobots may increase the availability in the long term [63].

If these systems are not available or invalid MEPs still occur, standardized ways to deal with coil corrections and invalid MEPs are needed. Many papers do not report these provisions, if applied at all [11,64,65], which complicates comparisons between dependent variables. In the present study, we addressed provisions by communicating the frequencies of invalid MEPs and coil corrections and by defining criteria for invalid MEPs.

A limiting factor to the results may be interactions between the sessions. In our case, we alternated between 1 and 20 Hz protocols which should evoke opposing neuromodulatory effects according to the lofi-hife heuristic. Therefore, interactions between these protocols like mutually extinguishing effects are possible.

In conclusion, the increase of objectivity and thus reliability of rTMS protocols is required to fully understand cortical excitability mechanisms and achieve validity. Our findings highlight the relevance of the reproducibility crisis in non-invasive brain stimulation techniques [66] by showing only low to moderate test-retest reliability and no clear evidence for the lofi-hife heuristic in 1 and 20 Hz rTMS protocols. These findings further underscore the relevance of uniform setups and transparent methodologies to reduce the sources of variability across experiments [23,24,44,47]. Valid results are more likely by increasing the reliability and the objectivity of rTMS and single pulse TMS. As the lofi-hife heuristic is based on research containing all this variability, it must be critically reevaluated. Sham-controlled investigations of the effects and reliability of low and high frequency rTMS using bigger datasets as well as longer observation periods are needed.

## Data Availability

The datasets generated during and/or analyzed during the current study are available from the corresponding author (CK) on reasonable request.

## Acknowledgments

The authors thank J. Mason for linguistic advice, P. Seidel for help with the setup and the volunteers who participated in this study and the test runs.

## Author contributions

MS, BL and SS contributed to study concept and design. KP did the data acquisition and figures. KP and MS processed the data as well as quality check control. CK, MS and KP conducted data analysis. CK, KP, MO, MS, SS and WM performed interpretation and drafted and revised the manuscript. KP, MS and MA took part in participants’ recruitment and selection process, clinical evaluation, and monitoring of participants. All authors reviewed the manuscript.

## Additional Information

### Competing Interests Statement

The authors declare that the research was conducted in the absence of any commercial or financial relationships that could be construed as a potential conflict of interest.

### Funding

Author SS received funding from the European Union’s Horizon 2020 Framework Programme (grant agreement number 848261, UNITI project) and CK, MO and SS received funding from dtec.bw – Digitalization and Technology Research Center of the Bundeswehr (MEXT project).

## Notes

### Competing Interest Statement

The authors have declared no competing interest.

### Author Declarations

Ethics committee of the University of Regensburg gave ethical approval for this work. (ethical approval number: 16-101-0305)

