## supplemental table 1 for "Limited evidence for validity and reliability of non-navigated low and high frequency rTMS over the motor cortex"

This Supplementary Data contains one table.

**Supplementary Table S1**

*Intraclass correlation coefficient (ICC) and Pearson’s r (r) between session 1 and 2*

Notes: Shown are the values for the total measurements as well as for the quarters: quarter 1 (Q1), quarter 2 (Q2), quarter 3 (Q3), quarter 4 (Q4). ICCs and *r* for the mean and median of the amplitudes of motor evoked potentials were calculated separately for the 20 Hz and 1 Hz protocols. Bold values mark moderate/medium reliability/effect size, other values are interpreted as markers of low reliability. CI = 95% confidence interval.

|  | **parameter** | **frequency** | **ICC** | **CI - ICC** | ***r*** | **CI - *r*** |
| --- | --- | --- | --- | --- | --- | --- |
| **whole** | **mean** | **1 Hz** | 0.281 | [-0.546, 0.662] | 0.162 | [-0.211, 0.493] |
|  |  | **20 Hz** | 0.204 | [-0.544, 0.605] | 0.123 | [-0.248, 0.463] |
|  | **median** | **1 Hz** | 0.480 | [-0.109, 0.755] | **0.322** | [-0.043, 0.611] |
|  |  | **20 Hz** | 0.162 | [-0.604, 0.582] | 0.097 | [-0.273, 0.442] |
| **Q1** | **mean** | **1 Hz** | 0.056 | [-1.05, 0.557] | 0.030 | [-0.334, 0.386] |
|  |  | **20 Hz** | 0.001 | [-0.789, 0.482] | 0.001 | [-0.359, 0.361] |
|  | **median** | **1 Hz** | 0.406 | [-0.26, 0.719] | 0.253 | [-0.118, 0.562] |
|  |  | **20 Hz** | -0.319 | [-1.389, 0.321] | -0.166 | [-0.497, 0.207] |
| **Q2** | **mean** | **1 Hz** | 0.087 | [-0.984, 0.572] | 0.044 | [-0.321, 0.398] |
|  |  | **20 Hz** | **0.519** | [0.043, 0.765] | **0.404** | [0.051, 0.667] |
|  | **median** | **1 Hz** | 0.356 | [-0.381, 0.697] | 0.215 | [-0.157, 0.534] |
|  |  | **20 Hz** | 0.394 | [-0.186, 0.701] | 0.267 | [-0.103, 0.572] |
| **Q3** | **mean** | **1 Hz** | 0.382 | [-0.327, 0.709] | 0.241 | [-0.131, 0.553] |
|  |  | **20 Hz** | 0.072 | [-0.683, 0.522] | 0.047 | [-0.319, 0.4] |
|  | **median** | **1 Hz** | 0.496 | [-0.073, 0.762] | **0.340** | [-0.023, 0.624] |
|  |  | **20 Hz** | 0.112 | [-0.7, 0.557] | 0.068 | [-0.3, 0.418] |
| **Q4** | **mean** | **1 Hz** | 0.469 | [-0.136, 0.75] | **0.344** | [-0.019, 0.627] |
|  |  | **20 Hz** | -0.242 | [-1.737, 0.421] | -0.151 | [-0.485, 0.221] |
|  | **median** | **1 Hz** | 0.411 | [-0.262, 0.722] | 0.293 | [-0.075, 0.591] |
|  |  | **20 Hz** | 0.152 | [-0.839, 0.602] | 0.123 | [-0.248, 0.463] |
